# Association between lipoprotein(a), *LPA* genetic risk score, aortic valve disease, and subsequent major adverse cardiovascular events

**DOI:** 10.1101/2023.09.20.23295872

**Authors:** Matthew K Moore, Gregory T Jones, Sally McCormick, Michael JA Williams, Sean Coffey

## Abstract

**Background:** Cohort studies have demonstrated associations between calcific aortic valve disease (CAVD) and Lp(a). As Lp(a) is almost entirely genetically determined, with the increasing clinical availability of genetic information, we aimed to determine if Lp(a), when predicted from genetic data, is associated with CAVD and major adverse cardiovascular events (MACE).

**Methods:** Patients undergoing coronary angiography were invited to participate in the study. Of 752 analysable participants, 446 had Lp(a) measured, and 703 had a calculable *LPA* genetic risk score (GRS). CAVD was categorized as absent/present and by severity. The primary outcomes were presence of CAVD at baseline, and MACE over seven years follow-up.

**Results:** The GRS explained 45% of the variation in Lp(a). After adjustment for cardiac risk factors and coronary artery disease, the odds of CAVD increased with Lp(a) (OR 1.039 per 10 unit increase, 95% C.I. 1.022 – 1.057, p<0.001) and GRS (OR 1.054 per 10-unit increase, 95% C.I. 1.024 – 1.086; p <0.001). Lp(a) and the GRS as continuous variables were not associated with subsequent MACE. Dichotomised GRS (>54) was associated with MACE, but this relationship became non-significant when coronary artery disease classification was added into the model (OR 1.333, 95% C.I. 0.927 – 1.912; p = 0.12).

**Conclusion:** An *LPA* GRS can explain 45% of the variation in Lp(a) levels, and both Lp(a) and the GRS are associated with CAVD. An elevated GRS is associated with future cardiac events in a secondary risk setting, but, if coronary artery disease status is known, it does not provide additional prognostic information.

## Introduction

Calcific aortic valve disease (CAVD) is a growing issue, with the number of non-rheumatic cases increasing by 76% in the past decade (1). Increasing age is the main risk factor for CAVD (2, 3), and despite the association of many other risk factors (4–7), there is no preventive or curative therapy aside from aortic valve replacement (AVR). Large cohort studies have demonstrated associations between high lipoprotein(a) (Lp(a)) and CAVD, both in terms of prevalence and incidence (8–11). Similarly, Lp(a) has been associated with major adverse cardiovascular events (MACE), in particular, nonfatal myocardial infarction, coronary death, and ischaemic stroke (12). A recent consensus statement from the European Atherosclerosis Society states that Lp(a) is a causal, continuous risk factor for CAVD, and recommends individuals have an Lp(a) measurement at least once in their lifetime (13). The vast majority of an individual’s Lp(a) level is genetically determined, and a recent publication by Burgess *et al* developed a genetic risk score (GRS) which predicted 50-60% of the variation in Lp(a) (a “genetically predicted” Lp(a) level) (14). This GRS has been previously associated with CAVD (15), but has yet to be tested in other populations. Although there has been significant interest garnered in what levels of Lp(a) confer the most risk, there remains significant variation between Lp(a) assays, with different assays of the same Lp(a) sample varying by 8-22% (16). Given that Lp(a) levels in a population can span nearly three orders of magnitude (17) and recommendations for broad screening, variations between assays may lead to imprecision in the estimation of risk.

This study aimed to determine if Lp(a), as either direct plasma protein levels or estimated using a GRS, is independently associated with CAVD or MACE in a New Zealand based clinical cohort.

## Methods

### Study participants and setting

The data for this study were obtained from consecutive individuals undergoing coronary angiography between January 2012 and May 2013 at Dunedin Hospital, New Zealand, who took part in a study examining the prevalence of abdominal aortic aneurysm in this patient population. All participants provided informed consent before their angiography, and ethical approval was given by the Upper South A Regional Ethics Committee on 26 January 2012 (Ref: URA/11/11/072). Of 1100 patients invited into the study, 48 declined involvement or were too unwell to participate. 1052 people (participation rate of 95.6%) thus comprised the primary study cohort. Further information on this cohort can be found in a paper by Jones *et al* (18). For this study, those without a baseline echocardiogram were excluded, giving a final study cohort of 783 people.

### Variables and potential confounders

Study participants filled out a questionnaire at baseline which detailed potential confounders. The following variables were collected by the study team at time of enrolment: age, sex, ethnicity, height, weight, waist-hip ratio, atrial fibrillation on ECG, previous percutaneous coronary intervention, history of previous disease (arthritis, dyslipidaemia, stroke, diabetes, hypertension), pack-years smoking, smoking status (current, ex, never), peripheral vascular disease, and chronic kidney disease. Indication for angiography, and overall coronary artery disease classification (using the criteria described in Jones *et al* (18)) were obtained from the electronic medical record.

### Follow up data collection

Participants were followed-up via their electronic health records (HealthConnectSouth, Orion Health, Auckland, New Zealand) to January 2021.

### Classification of calcific aortic valve disease

The presence of CAVD was determined by the clinical classification on the participants’ baseline echocardiography report and classified as either Prevalent CAVD (any disease, from sclerosis to severely calcified) or No CAVD. The prevalent CAVD group was further subdivided into the different strata of valvular disease: Early (sclerosis, mild, mild to moderate) or Late (moderate, moderate to severe, severe).

### Definition of major adverse cardiovascular events (MACE)

For this study, MACE is defined according to the American College of Cardiology/American Heart Association Key Data Elements and Definitions for Cardiovascular Endpoint Events in Clinical Trials (19). It is a composite outcome of stroke, acute myocardial infarction, hospitalization due to heart failure, hospitalization due to unstable angina, unplanned revascularization, and cardiovascular death. If a diagnosis of any MACE was recorded on a participant’s discharge summary, the date and type of MACE was recorded. In the instance of acute myocardial infarction, the type (STEMI/NSTEMI) was also recorded.

### Outcome measures

The primary outcomes were presence of calcific aortic valve disease, and MACE during the follow up period. Secondary outcomes were the subdivisions of CAVD (None, early, late) and the Individual components of MACE.

### Measurement of plasma Lp(a)

Plasma lipoprotein(a) concentrations (in nmol/L) were previously acquired for a subset of this cohort (n= 446 subjects that had indications for angiography related to myocardial infarction or unstable angina) as part of a study examining DNA methylation and Lp(a) (20). The specific enzyme-linked immunosorbent assay (ELISA)-based method used was described in that paper (20). A key element of this assay is that the antibodies used do not bind to the variable repeat regions of the apo(a) protein and the resulting Lp(a) measurements are therefore independent of apo(a) isoform size.

### Genetic risk score calculation

Burgess *et al* have previously developed a genetic risk score (GRS) that aimed to predict Lp(a) concentration using 43 single nucleotide polymorphisms (SNPs) in the *LPA* gene (14). The GRS genetically estimates the Lp(a) level for an individual. A subset of study participants (n=727) had whole-genome SNP data available, including a variety of SNPs of the *LPA* gene (20). In all, 40 (directly genotyped or imputed with high (>0.9) quality scores) of the 43 Burgess GRS SNPs were available for the Lp(a) GRS calculation. The GRS was calculated as:

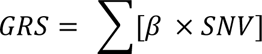

Where β is the conditional association with Lp(a), and SNV is number of effect alleles at that locus (0, 1 or 2). The units of the GRS were multiplied by 2.15 to convert from mg/dL to nmol/L, as has been done previously (21). A full list of the used genetic variants and their conditional associations with Lp(a) can be found in in the supplementary material (Table S1).

### Statistical methods

Data linked to a participant’s unique National Health Index (NHI) number was stored and managed securely using REDCap electronic data capture tools hosted at the University of Otago (22, 23). All analysis was performed on a de-identified dataset with the NHI numbers replaced with an anonymous identifier. Continuous data are expressed as mean (standard deviation) if normally distributed, and as median (interquartile range) if non-normally distributed. Data were analysed with the Mann-Whitney *U*-test if continuous and non-normally distributed, and with ANOVA if continuous and normally distributed. Categorical variables were analysed by the Chi-square test.

Lp(a) and GRS were analysed as both continuous and categorical variables. To determine dichotomised values between “high” and “low” Lp(a), receiver operating characteristic (ROC) curve analyses were conducted, and the Youden’s index used to identify associated criterion. MACE at seven years was treated as a binary variable. Kaplan-Meier survival curves were also produced, stratified by measured Lp(a) or GRS classification. Group differences were assessed using the log-rank test. Associations between valve disease and Lp(a)/GRS were measured using forward stepwise logistic regression. All data analysis was performed using R, version 3.6.3, and the R packages Survminer and tidyverse (24–26).

## Results

### Baseline characteristics

The demographics of the study cohort can be found in Table 1. In all, 26 of the 783 participants either had an AVR at baseline or did not have CAVD severity reported, and were excluded from the study, giving a final cohort size of 757. Of these participants, 433 had an available Lp(a) assay result, and 703 had a calculable GRS. Participants were followed up to the 31^st^ of January 2021, giving a mean follow up time of 8.34 ± 0.38 years.

**Table 1:**
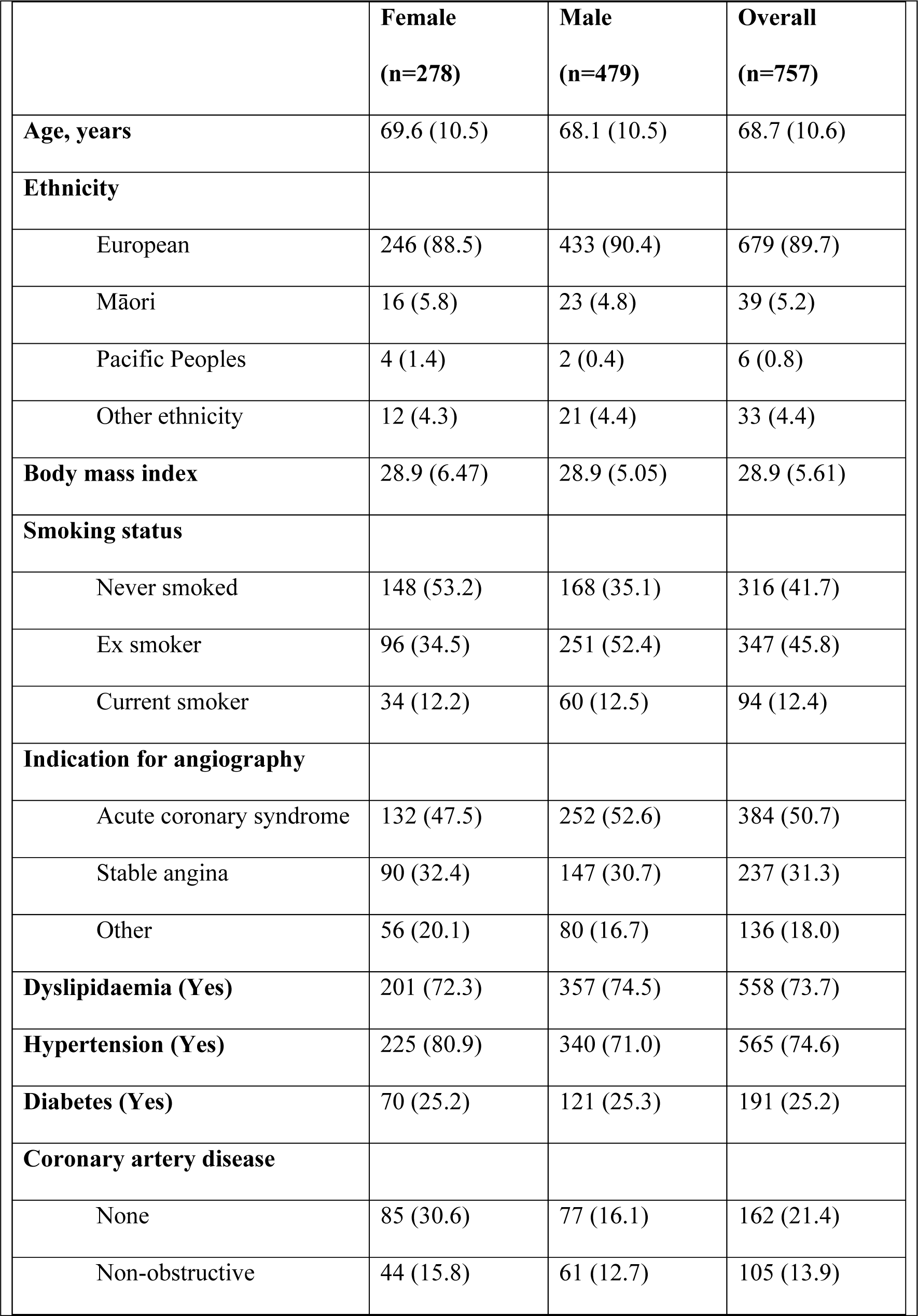
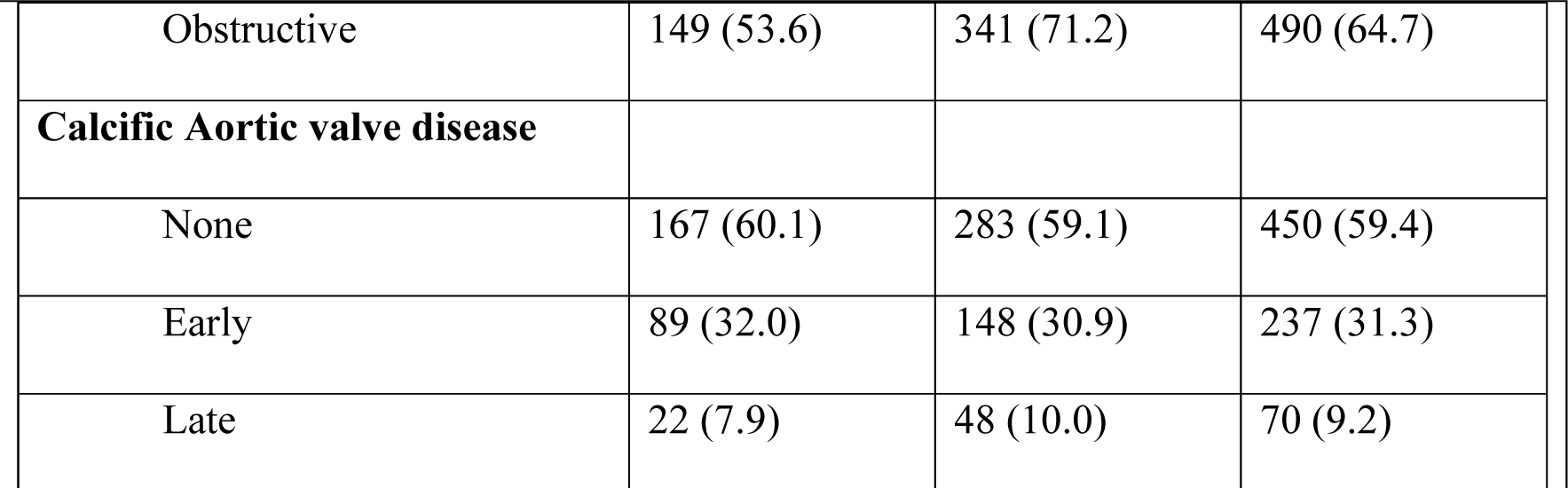
Demographics and clinical profile of the initial cohort with known native aortic valve status. Numeric variables presented as mean (standard deviation), categorical with percentages in brackets.

|  | **Female**<br>**(n=278)** | **Male**<br>**(n=479)** | **Overall**<br>**(n=757)** |
| --- | --- | --- | --- |
| **Age, years** | 69.6 (10.5) | 68.1 (10.5) | 68.7 (10.6) |
| **Ethnicity** |  |  |  |
| European | 246 (88.5) | 433 (90.4) | 679 (89.7) |
| Māori | 16 (5.8) | 23 (4.8) | 39 (5.2) |
| Pacific Peoples | 4 (1.4) | 2 (0.4) | 6 (0.8) |
| Other ethnicity | 12 (4.3) | 21 (4.4) | 33 (4.4) |
| **Body mass index** | 28.9 (6.47) | 28.9 (5.05) | 28.9 (5.61) |
| **Smoking status** |  |  |  |
| Never smoked | 148 (53.2) | 168 (35.1) | 316 (41.7) |
| Ex smoker | 96 (34.5) | 251 (52.4) | 347 (45.8) |
| Current smoker | 34 (12.2) | 60 (12.5) | 94 (12.4) |
| **Indication for angiography** |  |  |  |
| Acute coronary syndrome | 132 (47.5) | 252 (52.6) | 384 (50.7) |
| Stable angina | 90 (32.4) | 147 (30.7) | 237 (31.3) |
| Other | 56 (20.1) | 80 (16.7) | 136 (18.0) |
| **Dyslipidaemia (Yes)** | 201 (72.3) | 357 (74.5) | 558 (73.7) |
| **Hypertension (Yes)** | 225 (80.9) | 340 (71.0) | 565 (74.6) |
| **Diabetes (Yes)** | 70 (25.2) | 121 (25.3) | 191 (25.2) |
| **Coronary artery disease** |  |  |  |
| None | 85 (30.6) | 77 (16.1) | 162 (21.4) |
| Non-obstructive | 44 (15.8) | 61 (12.7) | 105 (13.9) |
| Obstructive | 149 (53.6) | 341 (71.2) | 490 (64.7) |
| **Calcific Aortic valve disease** |  |  |  |
| None | 167 (60.1) | 283 (59.1) | 450 (59.4) |
| Early | 89 (32.0) | 148 (30.9) | 237 (31.3) |
| Late | 22 (7.9) | 48 (10.0) | 70 (9.2) |

Of the 433 with an Lp(a) measure available, there were 172 (38.6%) cases of CAVD. Of these 172, 156 were “early” disease and 16 were “late” disease. Of the 703 with a calculable GRS, there were 283 (38.9%) cases of CAVD. Of these, 220 were “early” disease and 63 were “late” disease.

### Distributions of measured and GRS calculated Lp(a) and their association

The distribution of plasma Lp(a) was positively skewed, with a long rightwards tail (**Figure 1**). The data range spans nearly three orders of magnitude, with a median of 62 nmol/L (IQR: 24-147 nmol/L).

**Figure 1:**
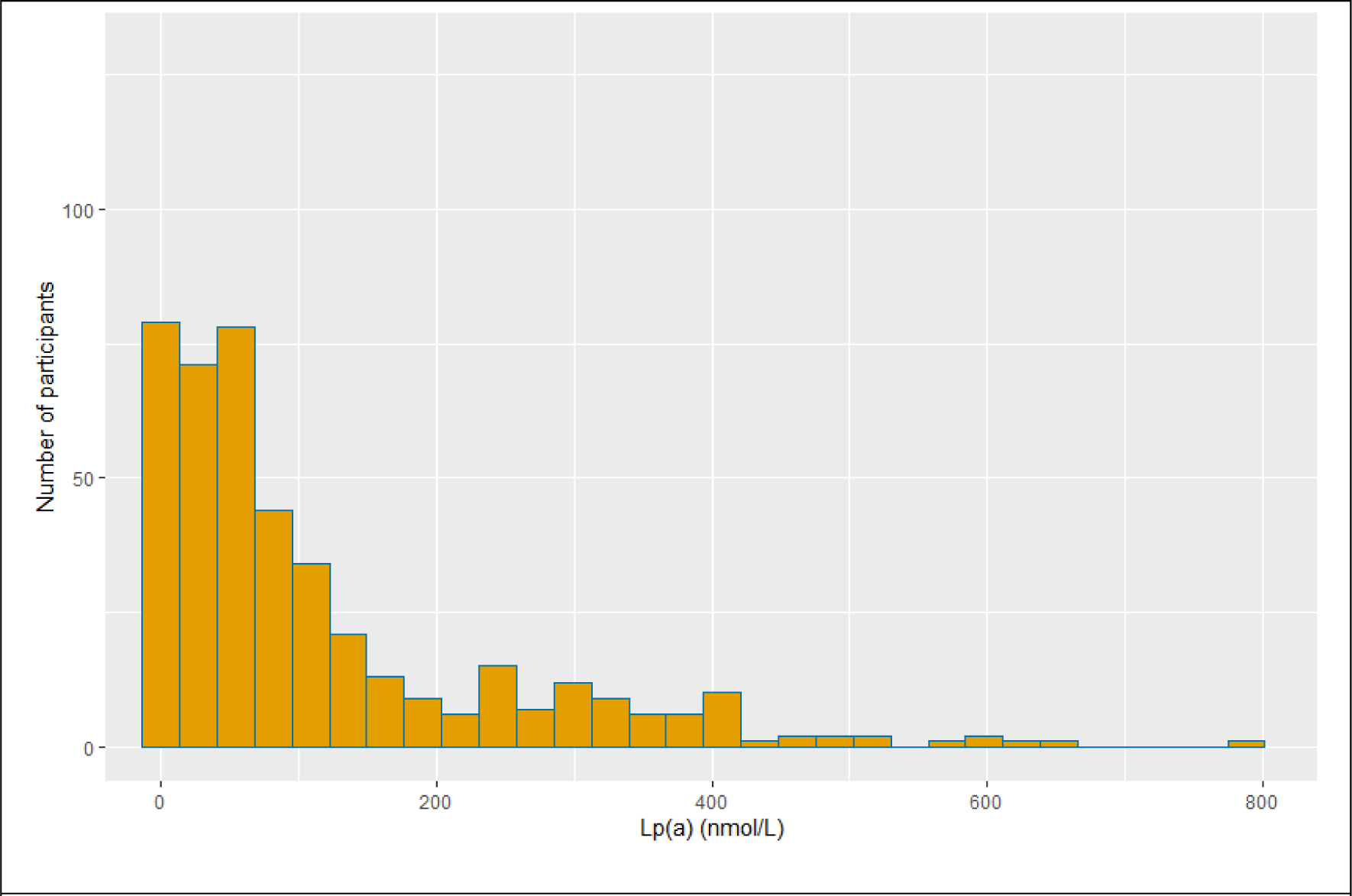
Distribution of measured plasma Lp(a) (nmol/L)

In the 703 participants with a calculable GRS, the distribution of the genetically predicted Lp(a) was bimodal (**Figure 2**) and had a smaller range with a median of 16.8 nmol/L (IQR: 2.47-65.90 nmol/L). It should be noted that an individual’s GRS could be below zero, as some of the conditional SNP associations were negative.

**Figure 2:**
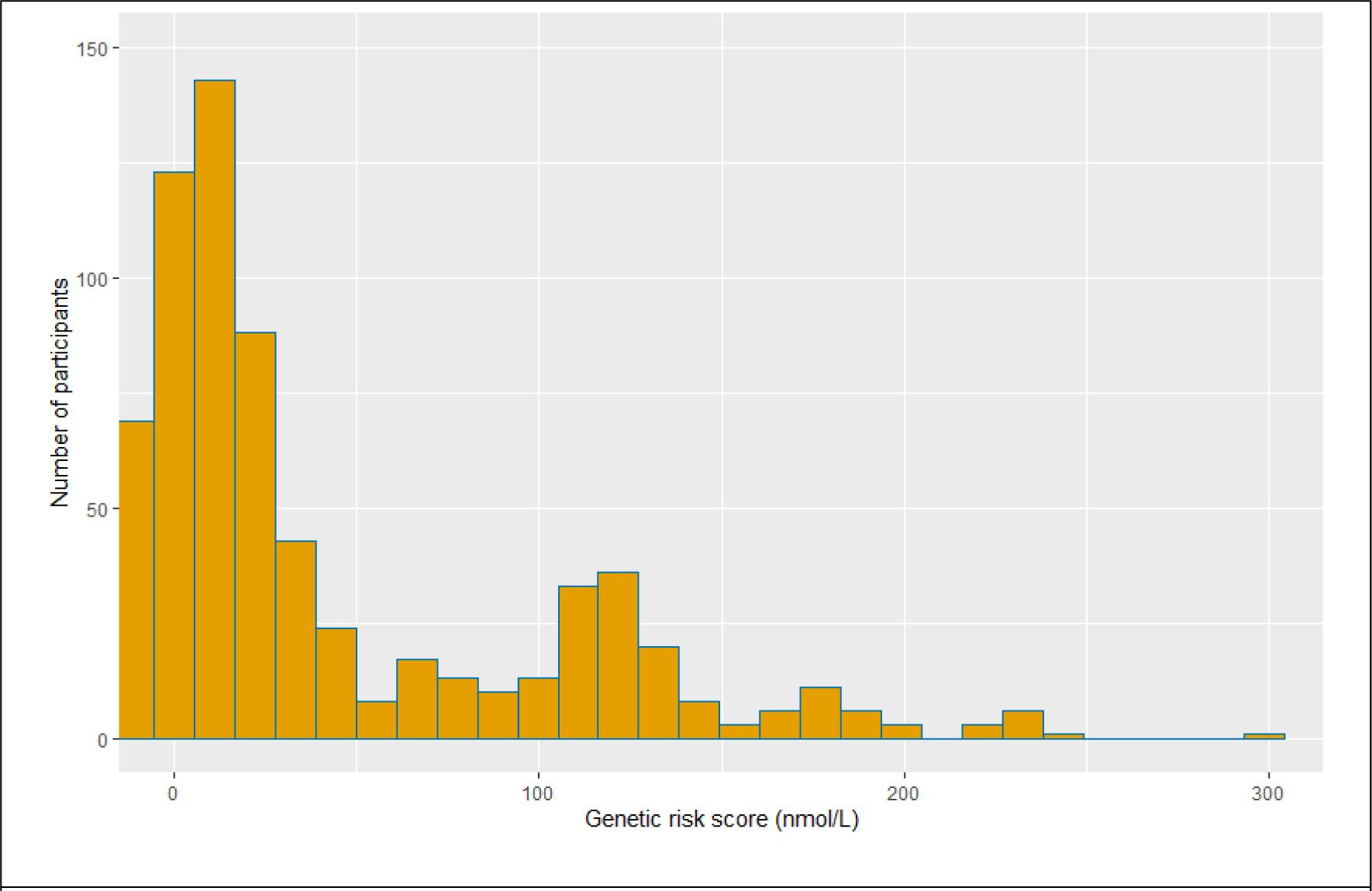
Distribution of genetic risk score (*genetically predicted* nmol/L)

In those with both GRS and measured Lp(a), measured plasma Lp(a) and the genetically predicted Lp(a) were significantly correlated (**Figure 3**; Measured Lp(a) = (GRS * 1.52) + 47.35, p < 0.001). The GRS appeared to explain 45% of the variation in plasma Lp(a) levels. A Bland-Altman plot of the GRS against measured Lp(a) is shown in **Figure 4**, showing a bias of −69.9, and uncertainty interval of −268.6 - 128.9 and increasing bias with increasing mean Lp(a) measurement.

**Figure 3:**
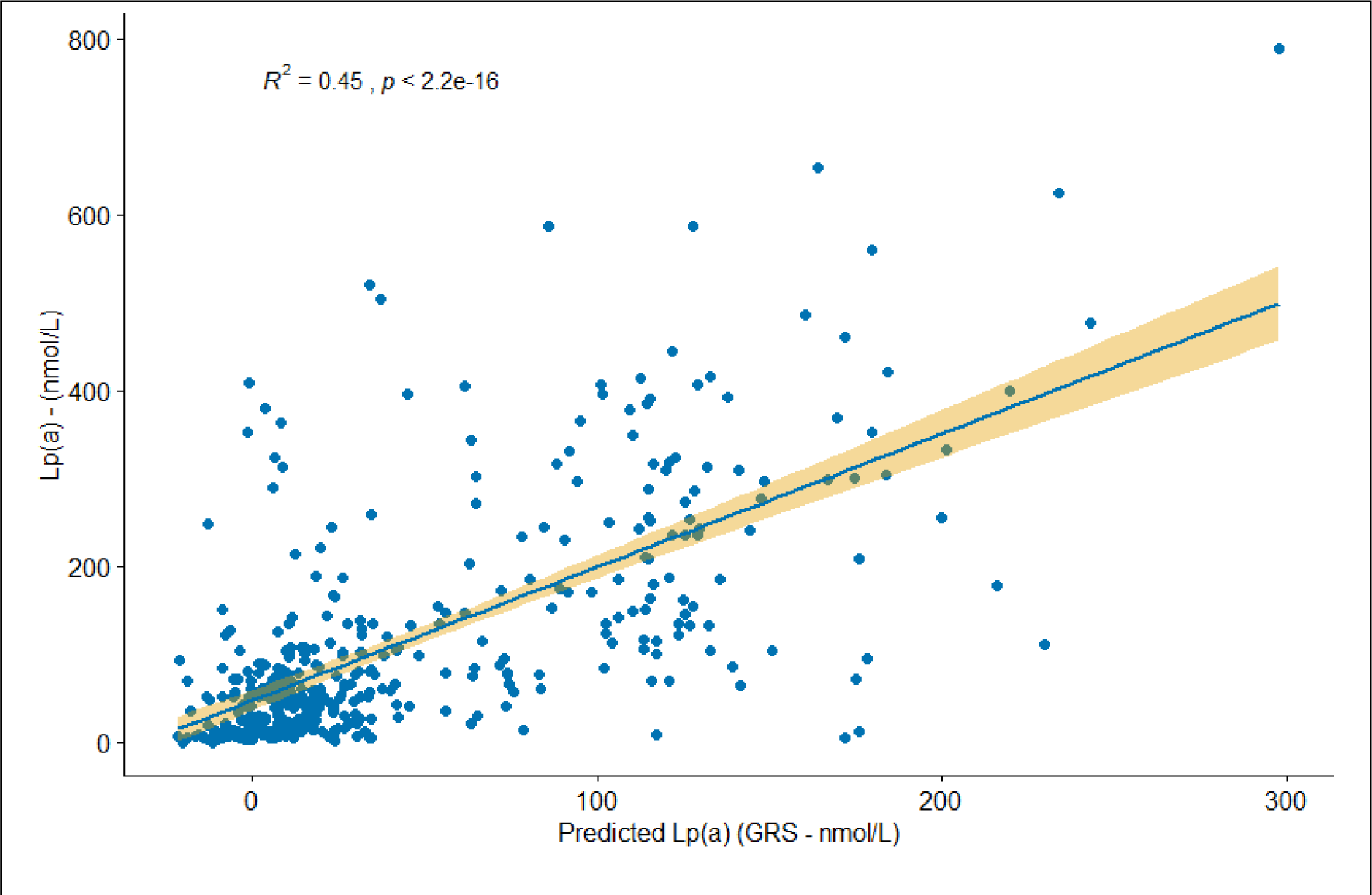
Scatterplot of the (GRS) genetically predicted Lp(a) vs measured plasma Lp(a)

**Figure 4:**
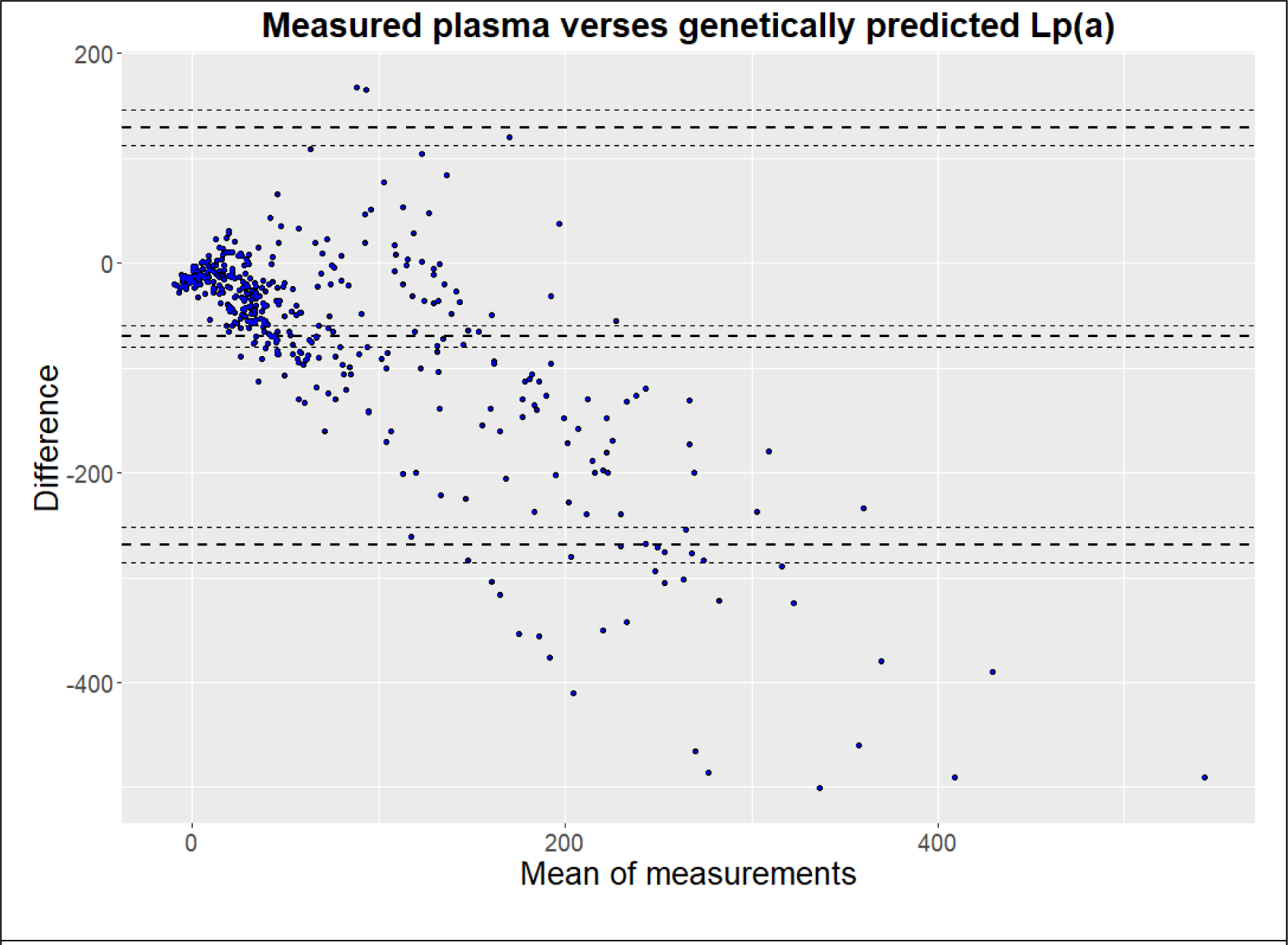
Bland-Altman plot of measured plasma Lp(a) versus genetic risk score. The calculated difference is measured GRS minus Lp(a).

### Association of measured Lp(a) with CAVD

A relatively small number of individuals had late CAVD, which made analysis across the strata of the secondary outcome (no CAVD versus early, or versus late) challenging. Only 16 participants had late CAVD in the plasma Lp(a) group, and 63 in the GRS group. As a result, comparisons are focused on no CAVD versus any CAVD.

Plasma Lp(a) as a continuous variable was significantly associated with CAVD in both the univariable and multivariable logistic regression analyses. A 10 nmol/L increase in Lp(a) was associated with 1.04 times the odds of having CAVD in the second adjusted model (see Table 2). The percentile plot of measured plasma Lp(a) levels (**Figure 6**) showed a divergence in levels between those with and without CAVD at the 50^th^ percentile, with the 75^th^ percentile (a commonly applied risk association cut-off) corresponding to an Lp(a) concentration of 147 nmol/L). Similarly, when examining the GRS percentiles plot (Figure S1), there was a divergence at the 60^th^ percentile, with the 75^th^ percentile corresponding to a predicted Lp(a) of 66 nmol/L.

**Figure 5:**
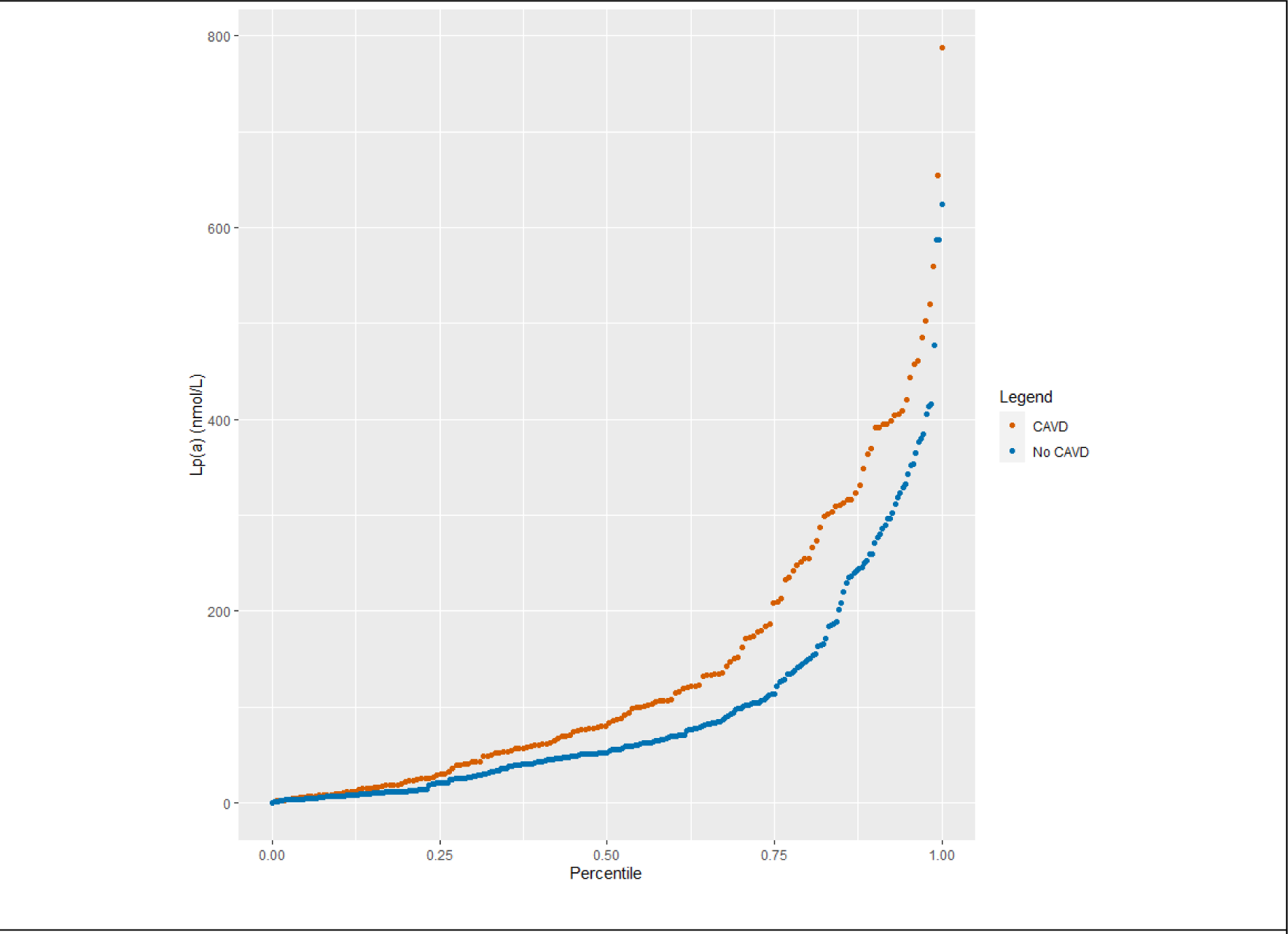
Percentiles plot of measured plasma Lp(a) stratified by CAVD status

**Figure 6:**
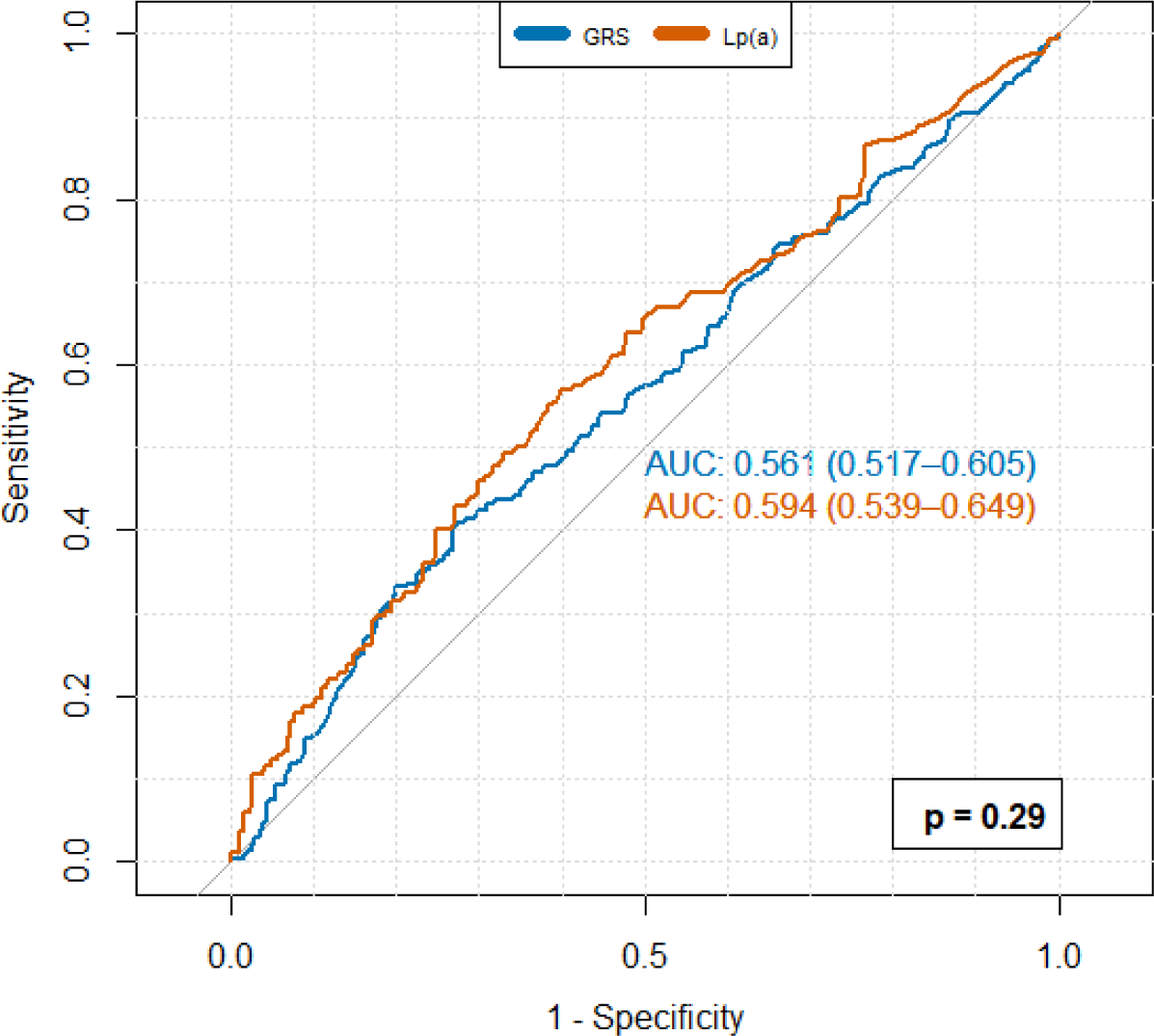
Receiver operating characteristic (ROC) curves for respective plasma Lp(a) and the GRS associations with (any) CAVD status. The difference between the two Lp(a) values was not significant (DeLong test p-value 0.29).

**Table 2:** Logistic regression model for (any) CAVD per 10 nmol/L increase in plasma Lp(a) * Adjusted for age, sex, BMI, dyslipidaemia, diabetes, hypertension, and smoking status ** Forward stepwise regression model adjusting for age, sex, BMI, dyslipidaemia, diabetes, hypertension, smoking status, stroke, and coronary disease classification

|  | **Odds ratio for plasma Lp(a)**<br>**per 10 nmol/L increase** | **P value** |
| --- | --- | --- |
| Univariate | 1.026 (95% CI 1.011-1.042) | <0.001 |
| Adjusted model 1* | 1.038 (95% CI 1.020-1.056) | <0.001 |
| Adjusted model 2** | 1.039 (95% CI 1.022-1.057) | <0.001 |

The ROC curve for plasma Lp(a) vs CAVD status showed area under the ROC curve of 0.59 (95% confidence interval 0.54-0.65, **Figure 6**). A cut-off value for “elevated” Lp(a) for further analysis was 69 nmol/L, calculated using the Youden’s index associated criterion of the ROC curve. At this level, there was a sensitivity 0.41 and specificity 0.43 for CAVD. Previous research has also identified the 75^th^ percentile as a high-risk value (2), and this was additionally investigated using multivariable logistic regression, with similar results (Table S2).

### Association of the genetic risk score (GRS) for Lp(a) with CAVD

Measured plasma Lp(a) and the GRS for Lp(a) were not significantly different from each other in their ability to predict prevalent CAVD (**Figure 6)**. For completeness, logistic regression was also performed on the GRS with CAVD as the binary outcome variable. A GRS ≥34.4 was identified as the optimal cut-off value for CAVD risk (sensitivity 0.28, specificity 0.59), using the Youden’s index associated criterion. In both the univariate and adjusted models, elevated GRS was associated with an increased odds of prevalent CAVD (**Table 3)**, however, the strength of this association was weaker than the models utilizing plasma Lp(a). The GRS as a continuous variable was also significantly associated with CAVD in the adjusted (age, sex, BMI, dyslipidaemia, diabetes, hypertension, smoking) model, with an odds ratio of 1.054 per 10 unit increase (95% C.I. 1.024 to 1.086; p < 0.001).

**Table 3:** Logistic regression model for CAVD, for GRS greater than 34.4. *Adjusted for age, sex, BMI, dyslipidaemia, diabetes, hypertension, and smoking status **Forward stepwise regression model adjusting for age, sex, BMI, dyslipidaemia, diabetes, hypertension, smoking status, stroke, and coronary disease classification

|  | **Odds ratio for GRS $\geq 34.4$** | **P value** |
| --- | --- | --- |
| Univariate | 1.778 (95% CI 1.293-2.446) | <0.001 |
| Adjusted model 1* | 1.963 (95%CI 1.380-2.803) | <0.001 |
| Adjusted model 2** | 2.002 (95% CI 1.402-2.869) | <0.001 |

### Association of Lp(a) and the GRS with MACE

For the MACE association analysis, those participants who were previously excluded for missing CAVD data, were reintroduced to the cohort. Over the follow-up period, 266 (34.0%) of the participants had a MACE. This represented an annualized event rate of 4.1%. Given that the shortest length of follow up was 7.6 years, the outcome variable of interest was seven year MACE.

Neither measured plasma Lp(a) or the Lp(a) GRS were associated with MACE at seven years using logistic regression (p = 0.88 for plasma Lp(a), p = 0.35 for Lp(a) GRS). However, using the optimal (elevated) GRS for predicting seven-year MACE (Youden’s index associated criterion value of 54nmol/L), suggested a significant association with shorter event-free survival (**Figure 7**– log-rank test p < 0.05).

**Figure 7:**
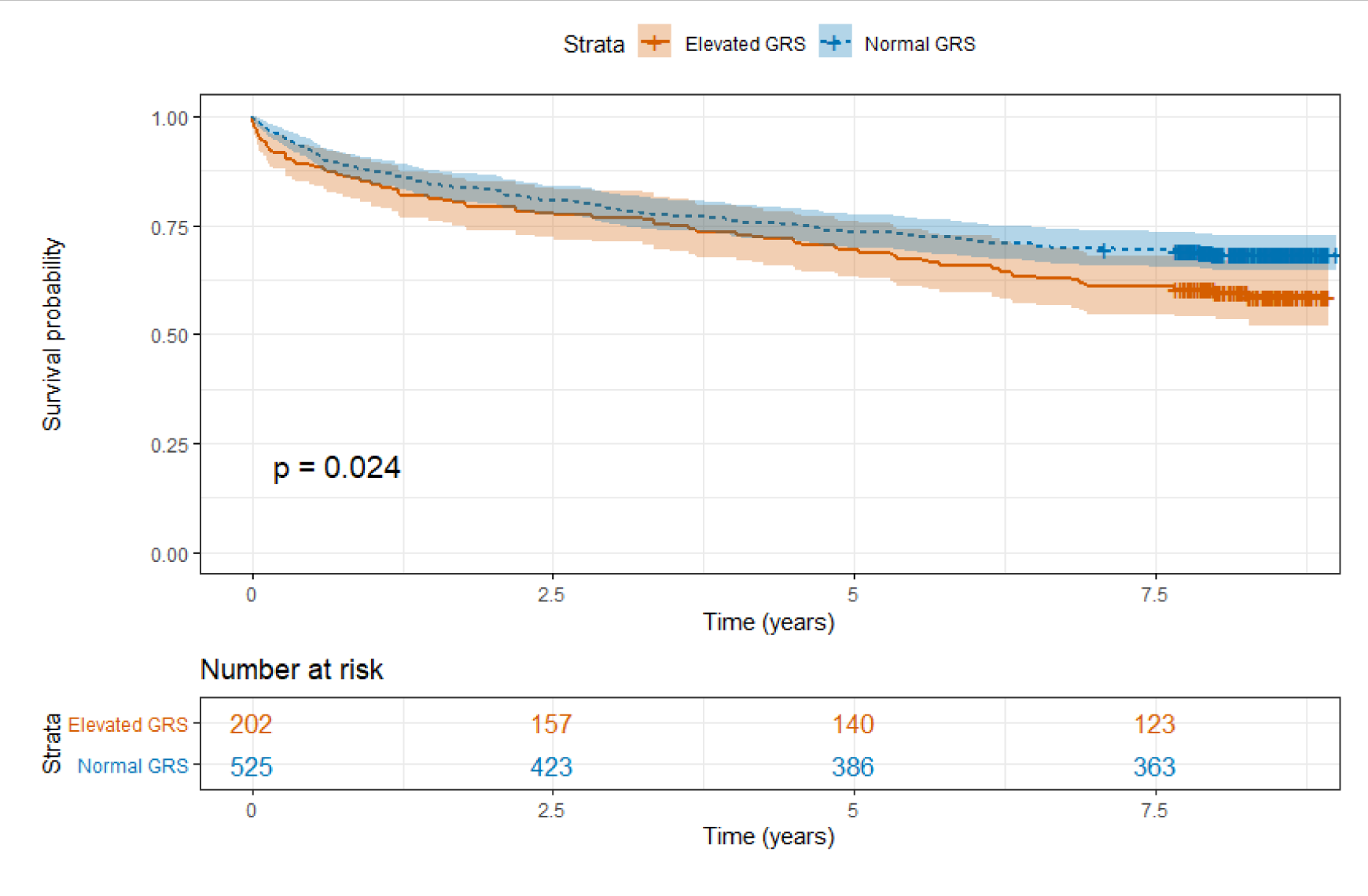
Kaplan-Meier survival curve with MACE as the outcome, stratified by GRS

**Figure 8:**
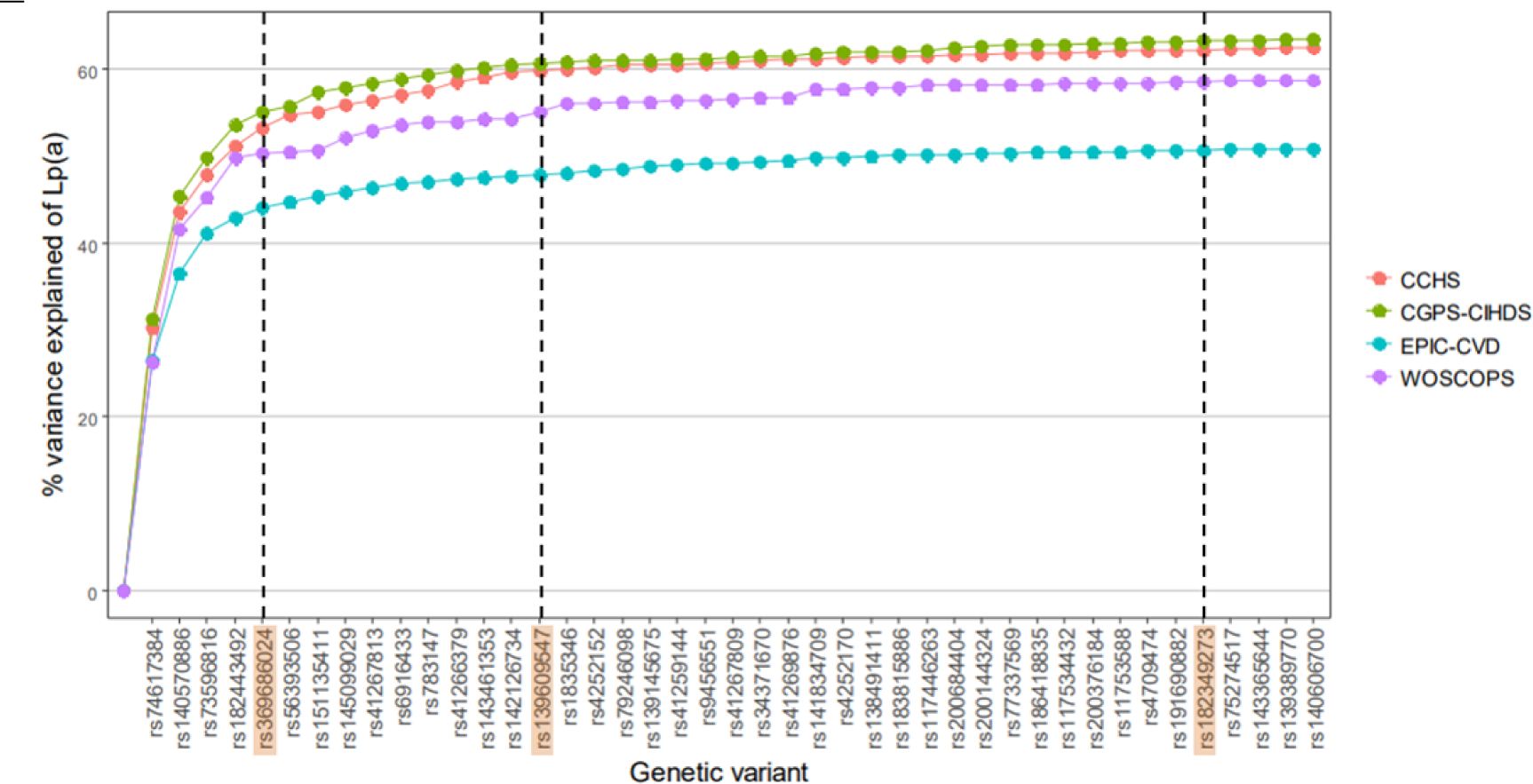
Variance of Lp(a) explained by each additional variant in each study. Modified from eFigure 5, Burgess *et al* 2018, JAMA Cardiol. (14) Highlighted SNPs were absent from our cohort.

Logistic regression analysis of the dichotomised GRS, using a cut-off value of 54, yielded a small but significant relationship with seven-year MACE (**Table 4Table**). However, this relationship was not significant if the status of coronary artery disease was added to the model.

**Table 4:** Logistic regression model for seven-year MACE, for GRS greater than 54. *Adjusted – age, sex, BMI, dyslipidaemia, diabetes, hypertension, and smoking status **Adjusted – as above + coronary disease classification

|  | **Odds ratio for GRS $\geq 54$** | **P value** |
| --- | --- | --- |
| Univariate | 1.478 (95% CI 1.044-2.121) | 0.024 |
| Adjusted model 1* | 1.489 (95% CI 1.044-2.121) | 0.027 |
| Adjusted model 2** | 1.333 (95% CI 0.927-1.912) | 0.119 |

In univariate analysis, dyslipidaemia (OR 2.07, p < 0.001), hypertension (OR 1.45, p < 0.05), being an ex-smoker (OR 1.51, p < 0.05), diabetes (OR 2.66, p < 0.001), BMI (OR per 1 unit increase 1.03, p < 0.05), non-obstructive CAD (OR 2.42 compared to no coronary artery disease, p < 0.01), and obstructive CAD (OR 3.94 compared to no CAD, p < 0.001) were associated with seven-year MACE. Current smoking was not associated with seven year MACE in this cohort.

## Discussion

Lp(a) has elicited significant research interest in the past two decades, largely due to its consistent associations with cardiovascular events and coronary disease and the development of Lp(a)-lowering agents currently in phase III clinical trial i.e. pelacarsen (27). Such interest has expanded into the field of CAVD. Associations between Lp(a) and CAVD vary depending on the population studied, if it is incident or prevalent disease as the outcome of interest, and whether Lp(a) is analysed as a continuous or binary outcome, or by percentiles (2, 28, 29). There is no clear consensus regarding the most appropriate “high-risk” threshold values, with in-sample 75^th^ or 80^th^ percentile values often being used, making comparison between studies and especially assessment of predictive validity difficult (30, 31). This may be due to previous mass and molar concentration measures that are the consequence of differing Lp(a) isoform sizes between individuals (32). Nevertheless, previous studies have shown a clear relationship between Lp(a) and both prevalent and incident CAVD. The Cardiovascular Health Study was the first large-scale study to investigate this association, showing that those with values above the 75^th^ percentile had a (adjusted) 23% greater risk of CAVD than those in the lowest quartile (2). Recent work in the Multi-Ethnic Study of Atherosclerosis and by Kaiser *et al* have found Lp(a) to be associated with aortic valve calcium, as measured by Agatson score on CT (28, 29). Several population studies have shown associations between elevated Lp(a) and incident CAVD (8, 10, 11). The European Atherosclerosis Society notes Lp(a) to be a continuous, causal risk factor for CAVD, based on the strong genetic association between the *LPA* gene and CAVD (13). Our present study aimed to evaluate both direct and genetically predicted measures of Lp(a) as a risk factor for prevalent CAVD and MACE in a New Zealand cohort.

Plasma Lp(a) was significantly associated with CAVD in both continuous and categorical terms (OR 1.04 per 10 nmol/L increase (95% C.I. 1.02 – 1.06)), an observation which is consistent with the previous literature (2, 28, 29). Interestingly, adjustment for confounding variables increased the strength of the relationship, potentially suggesting that cardiovascular risk factors may be causing a bias towards the null value. Almost the entire cohort had a calculable Lp(a) genetic risk score, allowing for a greater capture of the characteristics of such a secondary risk population.

In an analysis of the ROC curves comparing the GRS to plasma Lp(a), there was no significant difference in the predictive power for prevalent CAVD, which was relatively limited on an individual basis. The GRS is still relatively novel and our study is, to our knowledge, only the second study to validate its use for predicting CAVD (15). Utilization of genetic data instead of the Lp(a) assay can be advantageous by reducing measurement error and variation between studies due to the assay used, rather than characteristics of the cohort driving variation. Though we note our GRS performed worse than other studies for predicting “true” Lp(a); 45% of the variation was captured by the GRS, compared with 53-61% in other cohorts (4, 12), there was no significant difference between Lp(a) and the GRS for predicting prevalent CAVD. This suggests that although the GRS predicted only 45% of the variation in measured Lp(a), it shows promise as a surrogate for Lp(a) population analysis. The Bland-Altman plot (**Figure 4**) suggests that GRS is unlikely to be accurate enough to predict measured Lp(a) on an individual basis. However, further variants associated with Lp(a) concentration have been identified, which presents the possibility to iteratively improve the GRS (33).

Our study also examined if plasma Lp(a) or a Lp(a) GRS could be used to predict MACE at seven years follow-up. Plasma Lp(a) was not associated with either seven year incident MACE as a binary outcome or time to incident MACE. This conflicts with the results of other studies in this field (12), and we suspect this is due to a degree of ascertainment bias; as the population assessed was a group with a relatively high burden of cardiovascular disease, the information conveyed by Lp(a) might have been attenuated by the information contained in coronary artery assessment. Although a GRS cut-off value of 54.4 was associated with seven year MACE and time to MACE, this association was small and became insignificant once coronary disease status was taken into account. It was also not associated with seven year MACE or time to MACE as a continuous variable. This is to say that, if the coronary disease status is known, the GRS does not appear to provide any additional information for risk prediction. If the coronary disease status is unknown, the GRS may represent a statistically significant risk estimate, however, due to its small effect size it appears unlikely to be a clinically useful MACE risk marker. This suggests that the effect of Lp(a) on MACE is primarily mediated through its effect on incident coronary artery disease.

There are several strengths to our study. Firstly, this cohort had a very high participation rate and a very low loss to follow-up. Secondly, the plasma Lp(a) assay used was a isoform size-insensitive molar assay, which is more likely to accurately reflect Lp(a) levels than other size-dependent assays. Thirdly, New Zealand has a well-established public healthcare system with very few procedures or events occurring in the private setting, meaning that there is a low chance of unrecorded events during follow-up.

There is now a small but consistently statistically significant body of work associating elevated circulating Lp(a) with prevalent CAVD, with several studies also supporting a relationship with incident CAVD. The predicted increase in CAVD burden globally (34), given that the only effective treatment is valve replacement late in the disease process, means that a search for preventative or mitigatory therapies is paramount. Lp(a) has been identified as a treatment target for the prevention of cardiovascular events in several clinical trials (35–38), and it is plausible that it could be a target for the prevention of CAVD as well, given its implication in the pathophysiology of CAVD (39). However, no trial has been undertaken to evaluate if Lp(a) lowering therapies could reduce the risk of incident CAVD, and such a trial would be costly and require lengthy follow-up times. The disease process for CAVD is slow, beginning with sclerotic valvular disease that may take many decades before progression to calcification. It is unclear what proportion of those with sclerosis will go on to develop aortic stenosis, and over what time frame. As discussed in the background, we have only been able to identify two studies that examine the progression of sclerosis to more advanced calcification (40, 41). Furthermore, most cases of CAVD are asymptomatic, and such a trial would require regular echocardiographic screening of all participants. Coupled with the relatively low incidence rates of CAVD in the general population (5, 42–44), such a trial would be difficult to accomplish.

Our study is not without limitations, however. As previously discussed, this was a secondary risk cohort of participants undergoing coronary angiography, and the results here are not necessarily applicable to the general population. Our cohort had 40 of the 43 SNPs used for the original genetic risk score by Burgess *et al,* but we do not expect this to appreciably alter the final score in our cohort. The three absent SNPs would likely explain only an additional 1-2% of variation (Figure 12), which would be unlikely to significantly alter our results.

## Conclusion

An *LPA* genetic risk score can explain 45% of the variation in lipoprotein(a) levels. Both Lp(a) and an *LPA* genetic risk score are associated with the presence of CAVD. An elevated GRS appears to be associated with future cardiac events in a secondary risk setting, but if the coronary artery disease status is known it does not appear to provide additional prognostic information. The GRS may be useful in evaluating population-level associations between Lp(a) and cardiovascular and valve-related outcomes, especially in groups where the coronary artery disease status is unknown.

## Data Availability

Due to the New Zealand National Ethical Standards, external data sharing is not permitted.

## Supplemental material (see separate file)

Tables S1-S2

Figure S1

